# Neutrophil extracellular traps linked to idiopathic pulmonary fibrosis severity and survival

**DOI:** 10.1101/2024.01.24.24301742

**Authors:** Scott M. Matson, Linh T. Ngo, Yui Sugawara, Veani Fernando, Claudia Lugo, Imaan Azeem, Alexis Harrison, Alex Alsup, Emily Nissen, Devin Koestler, Michael P. Washburn, Michaella J. Rekowski, Paul J. Wolters, Joyce S. Lee, Joshua J. Solomon, M. Kristen Demoruelle

## Abstract

**Background:** Idiopathic pulmonary fibrosis (IPF) leads to progressive loss of lung function and mortality. Understanding mechanisms and markers of lung injury in IPF is paramount to improving outcomes for these patients. Despite the lack of systemic involvement in IPF, many analyses focus on identifying *circulating* prognostic markers. Using a proteomic discovery method followed by ELISA validation in multiple IPF lung compartments and cohorts we explored novel markers of IPF survival.

**Methods:** In our discovery analysis, agnostic label-free quantitative proteomics differentiated lung tissue protein expression based on survival trajectory (n=10). Following selection of the candidate pathway (neutrophil extracellular trap (NET) formation), we subsequently validated the presence of NETs in the IPF lung microenvironment using fully quantitative assays of known NET remnants in separate IPF cohorts (n=156 and n=52) with bronchoalveolar lavage fluid. We then assessed the correlation of these markers with baseline pulmonary function and survival.

**Results:** Discovery lung tissue proteomics identified NET formation as significantly associated with poor IPF survival. Using fully quantitative confirmatory tests for reproducibility we confirmed the presence of NET markers in IPF BALF and found significant correlations with worse pulmonary function in both cohorts (p<0.03 and p = 0.04 respectively). In the survival cohort, higher levels of NET markers predicted worse survival after adjusting for gender, age, and baseline physiologic severity (hazard ratio range: 1.79–2.19).

**Conclusions:** NET markers were associated with disease severity and worse survival in IPF. These findings suggest NET formation contributes to lung injury and decreased survival in IPF and may represent a potential therapeutic target.

## Background

Idiopathic pulmonary fibrosis (IPF) is a devastating and universally progressive condition with survival of 3 to 5 years from the time of diagnosis [1]. Current treatment options with antifibrotic therapy have been shown to slow lung function decline compared to placebo but fail to improve survival [2, 3]. Given the limitations of currently available therapies and the grim prognosis of IPF, the discovery of novel mechanisms of lung injury and targetable endotypes of IPF could offer new therapeutic targets.

Agnostic, systems biology methods, such as label-free quantitative proteomics, have been explored in IPF with a focus to identify proteomic signatures that distinguish IPF from normal control populations [4–10]. These findings have led to important insights into IPF pathobiology, but key features associated with survival in IPF remain elusive. Despite the median survival rate being dismal, there exists a spectrum of survival across all IPF patients. Therefore, we leveraged IPF patients with distinct survival trajectories and applied a shotgun, agnostic proteomic method in lung tissue to compare differential protein expression and pathways that could identify novel candidate targets associated with poor IPF survival. We then confirmed tissue proteomics findings in the IPF lung microenvironment via absolute quantification of pathway-associated protein targets in banked bronchoalveolar lavage fluid (BALF) from two independent IPF cohorts and assessed the clinical impact of these pathway-associated markers.

## Methods

### Study participants - Lung tissue proteomics cohort

We obtained surgical lung biopsy tissue samples from the interstitial lung disease (ILD) biorepository at National Jewish Health (NJH) that included patients with ILD who were prospectively enrolled into the Specialized Center of Research Study. A subset of these patients have been previously reported on [11]. We included 10 patients who had undergone prospective research lung biopsy who met the current IPF diagnostic criteria [12], which was confirmed by contemporary chart review (JJS, MKD). To best distinguish features associated with poor survival, two matched groups of five patients each were selected based on differential survival from the time of biopsy matched for age and baseline severity as determined by FVC% at the time of surgery (Table 1). All tissue samples underwent label-free quantitative proteomics, as described below. All samples were collected between 1986 to 2009. Clinical and demographic information was extracted from the medical record to include demographics, medications, and pulmonary function testing (PFT) results at the time of tissue collection (+/- 90 days). Date of death was confirmed using the United States Center for Disease Control’s National Death Index (CDC NDI).

**Table 1.** IPF Lung tissue discovery cohort.

|  | *Short Survival*<br>*(n=5)* | *Long Survival*<br>*(n=5)* | *P-value* |
| --- | --- | --- | --- |
| Age, mean (SD) | 63 (10.7) | 61.6 (6.1) | 0.81 |
| Gender, n (%) male | 4 (80%) | 2 (40%) | 0.41 |
| Smoker, n (%) ever | 4 (80%) | 2 (40%) | 0.41 |
| FVC% predicted, mean (SD) | 50.6 (13.5) | 67 (12.9) | 0.086 |
| DLCO% predicted, mean (SD) | 73.6 (21.3) | 92.5 (6.8) | 0.14 |
| On immunosuppression, n (%) | 2 (40%) | 0 (0%) | - |
| Survived to 5-years, % | 0 (0%) | 1 (20%) | - |
| Time to death in days, mean (SD) | 143.2 (89.5) | 1394.8 (881.2) | < 0.01 |
| Time to death in years, mean (SD) | 0.4 (0.3) | 3.82 (2.4) | < 0.01 |
| Protein count, mean (SD) | 1152 (262) | 1146 (130) | 0.90 |

### Study participants – Survival confirmation cohort

The above mentioned ILD NJH biorepository also obtained and stored research BALF from patients with IPF. For a survival confirmation analysis, we included all IPF patients in the ILD NJH biorepository with adequate BALF volumes available (N=156). This cohort was chosen for survival confirmation because given the timeframe of sample collection (1986 to 2009), there was nearly complete survival data for each participant which allowed for a robust survival analysis of any candidate target. As described above, all subjects met current IPF diagnostic criteria (confirmed by contemporary chart review), clinical and demographic information was extracted from the medical record, and date of death was confirmed using the CDC NDI.

### Study participants – Contemporary confirmation cohort

Given the era of collection of the survival confirmation cohort, a portion of IPF patients were on immunomodulatory therapy (29%) and none were on anti-fibrotic medications, which differs from the current clinical approaches to IPF management. To confirm contemporary applicability of the pathway of interest we included a contemporary IPF BALF confirmation cohort. This cohort included 27 IPF BALF samples from the University of California San Francisco (UCSF) ILD biorepository and 25 IPF BALF samples from the Weighing Risks and Benefits of Laparoscopic Anti-Reflux Surgery in Patients with IPF (WRAP-IPF) cohort [13]. UCSF samples were collected between 2013-2018 and WRAP-IPF samples were collected between 2014-2016. Clinical and PFT data were collected as part of the research study. Survival data was insufficient in this cohort, therefore, analyses with this cohort were limited to cross-sectional relationships with disease severity but provide an important assessment of relevance to contemporary IPF patients.

#### Lung tissue collection

During the initial prospective enrollment of the Specialized Center of Research Study at NJH, each patient underwent research protocol open thoracotomy or video-assisted thoracoscopic lung biopsy to confirm the usual interstitial pneumonia (UIP) pattern. These biopsies were taken from at least two separate sites in the same lung (upper and lower lobes) as previously reported [11]. Only subjects with banked lung tissue in adequate size and condition were considered for selection for the Lung tissue proteomics cohort.

#### BALF collection

BALF was collected using saline lavage. Cell count and differential were determined using an unspun BALF aliquot in the survival confirmation cohort. BALF cell-free supernatant was obtained following centrifugation and stored at -80^0^C until analysis.

#### PFTs

Pulmonary function testing was performed under normal conditions in routine clinical care, for prospective research purposes, or in the case of WRAP-IPF subjects, prior to surgical procedure. Percent predicted forced vital capacity (FVC%) and percent predicted diffusing capacity for carbon monoxide (DLCO%) at the time of BALF collection (+/- 3 months) were used to determine baseline disease severity.

#### Lung tissue protein isolation

Fresh frozen lung tissues were sliced, weighed, and thawed on ice. RIPA buffer spiked with protease inhibitors and phosphatase inhibitors was added to tissue samples (30µL RIPA/1mg tissue). The samples were then homogenized using BeatBox homogenizer (PreOmics) and manufacturer’s kit on standard power for 10 minutes twice. Tissue lysates were centrifuged at 14,000g for 10 minutes and supernatants free of cellular remnants and particulates were transferred to fresh tubes for protein isolation.

Total protein level for each tissue lysate was quantified using the Pierce BCA protein assay following manufacturer’s protocol against a standard curve of BSA. Standards and samples were measured with a plate reader at an absorbance of 562 nm. Proteins from tissue lysates were then isolated using acetone precipitation. An adequate volume of supernatant containing at least 50µg protein was aliquoted, reduced, alkylated, and proteins then precipitated with 3x volume ice cold acetone overnight at -20°C. The protein was pelleted and digested overnight with trypsin.

#### Label-free quantitative proteomics

Digested peptides from lung tissue were analyzed by nanoLC-MS/MS using a Vanquish Neo nano-UPLC interfaced directly to the Orbitrap Ascend Tribrid mass spectrometer (Thermo Fisher) equipped with a FAIMS source (details in Supplement Methods). Spectra were searched with Proteome Discoverer 3.0 against the human database downloaded from Uniprot on May 5, 2023, and a database of 155 common contaminants.

#### Neutrophil extracellular trap (NET) remnant testing

After completion of the proteomics analysis, the candidate pathway carried forward was neutrophil extracellular trap formation (NETosis). All BALF cell-free supernatant from the survival confirmation cohort was tested for levels of NET remnants using four complementary, but distinct, absolute quantitative methods [14–23]. These quantitative NET remnant assays included a research-based sandwich ELISA for MPO-DNA, an immunofluorescence assay for extracellular DNA (exDNA; Invitrogen Quanti-iT^TM^ PicoGreen® dsDNA assay), an ELISA for calprotectin (Werfen) and an ELISA for extracellular NE (exNE; Abcam). Given limitations of BALF volume in the contemporary confirmation cohort, only BALF exDNA was tested in this cohort as it required the smallest amount of BALF. For the commercially available assays, exDNA, calprotectin and exNE, manufacturers protocols for testing were followed. For MPO-DNA, previously published protocols were used [24]. Briefly, a high-binding EIA/RIA 96-well plate (Costar) was coated overnight at 4°C with anti-human MPO antibody (Bio-Rad0400-0002) in coating buffer from Cell Death Detection ELISA kit (Roche). The plate was then washed three times with 0.05% Tween 20 in PBS and blocked with 1% BSA in PBS for one hour at room temperature (RT). Following three more washes, BALF samples were diluted 1:2 in the blocking buffer and incubated for one hour at RT. The plate was then washed five times, followed by incubation with anti-DNA antibody for one hour at RT (HRP-conjugated, Cell Death kit, diluted 1:100 in blocking buffer). After 5 more washes, the plate was developed with 3,3’,5,5’-TMB substrate (Invitrogen) followed by a 2N sulfuric acid stop solution. Absorbance was measured at a wavelength of 450 nm. A background control well was run for each sample that included the BALF sample without including the primary anti-MPO antibody and background absorbance level was subtracted from each sample to account for any non-specific background binding of the sample.

### Statistical Analysis

#### Tissue proteomics data analysis

To identify discriminating lung tissue proteins between the short survival versus long survival groups, contaminant proteins and proteins detected in less than 50% of samples were excluded from analysis. Protein abundance data was normalized, log2 transformed, and median centered. For each protein, fold change (FC) and p-value between short survival and long survival was calculated. Proteins considered differentially expressed must have a p-value ≤ 0.05 and an absolute log_2_FC ≥ 1 (at least 2-fold change in either direction) for lung tissue data. Only differentially expressed proteins were utilized to perform an overrepresentation analysis (ORA) using QIAGEN Ingenuity Pathway Analysis (IPA) [25]. The Core Analysis workflow was used to generate the Canonical Pathways results.

Pathways were ranked based on p-value, and a pathway with an overlap p-value ≤ 0.05 was considered statistically significant. IPA also considers expression changes to predict pathway activation: A positive z-score suggests activation and a negative z-score suggests inhibition; however, a z-score = 0 or NaN (not a number) does not necessarily imply a prediction of no alternation. All z-scores were included in analysis per IPA recommendation.

#### Confirmation cohorts

Correlations between log transformed levels of NET remnants (DNA-MPO, exDNA, calprotectin, and exNE), BALF% neutrophils, FVC% predicted and DLCO% predicted were calculated using Pearson’s correlation coefficient. Linear regression models were used to compare FVC% predicted and DLCO% predicted with log transformed BALF MPO-DNA, exDNA, calprotectin, and exNE levels, sex, and age.

#### Validation survival analysis

In the survival confirmation cohort, Cox proportional-hazards (PH) models were used to assess the association between survival time (up to 5-years) and log transformed BALF MPO-DNA, exDNA, calprotectin and NE levels. Multivariate Cox PH models were used to determine associations with death adjusting for batch, GAP score [26], use of immunosuppressing medications (i.e., corticosteroids, mycophenolate mofetil, azathioprine, cyclophosphamide). We also dichotomized the survival confirmation cohort into high and low BALF NET remnant groups using a cut point function that determines the optimal point for discriminating between two groups for a continuous variable using the maximal log rank statistic using the surv_cutpoint() function in R, with a minimal proportion of observations per group set at 0.15. In this cut point analysis, we determined optimal levels for discriminating survival associated with each BALF NET remnant. Any patient with a BALF NET remnant level greater than the optimal cut point was categorized into the high group, and those with a level less than the cut point were categorized into the low group. Kaplan-Meier curves were used to visualize the survival function for high and low NET remnant groups. In all models, a p-value <0.05 was considered statistically significant.

All statistical analysis were performed in R version 4.1.3 unless specified otherwise.

## Results

### Patient characteristics

In the lung tissue proteomics cohort (n=10), the short survival group mean days to death was 143.2 (+/-89.5) and the long survival group mean days to death was 1394.8 (+/-881.2) (mean difference of 1251.6 days, p-value < 0.01). Age, FVC% and DLCO% were similar across the two groups, although the short survival group had a higher proportion of patients on immunosuppression (Table 1). Most patients in the cohort were male with a history of smoking.

In the survival confirmation cohort (n=156), the mean age was 66.1 years, and most patients were male (64.1%) ever-smokers (66.7%) (Table 2). The mean (SD) baseline % predicted FVC was 66.3 (17.5) and % predicted DLCO was 50.8 (16.0). Within the 5 years following BALF collection, 69.9% of participants had died. Twenty-nine percent of the cohort was taking immunosuppressing medications included prednisone, azathioprine, or mycophenolate mofetil at the time of BALF collection and none were on anti-fibrotic medications.

**Table 2.** Confirmation cohorts.

|  | ***Survival***<br>***Cohort***<br>***(n=156)*** | ***Contemporary***<br>***Cohort***<br>***(n=52)*** | ***P-value*** |
| --- | --- | --- | --- |
| Age, mean (SD) | 66.1 (7.6) | 68.7 (5.7) | 0.01 |
| Sex, n (% male) | 100 (64.1%) | 43 (82.7%) | 0.02 |
| Smoker, n (% ever) | 104 (66.7%) | 32 (61.5%) | 0.9 |
| FVC% predicted, mean (SD) | 66.3 (17.5) | 73.2 (16.6) | 0.01 |
| DLCO% predicted, mean (SD) | 50.8 (15.9) | 42.8 (11.9) | <0.01 |
| On immunosuppression, n (%) | 45 (28.9%) | 0 | - |
| On antifibrotic, n (%) | 0 | 22 (42.3%) | - |
| Survived up to 5-years, % | 30.1% | *17.3% | 0.10 |
| Time to death in days, mean (SD) | 1326.9 (1403.6) | 1269.0 (994.8) | 0.8 |
| Time to death in years, mean (SD) | 3.6 (3.8) | 3.5 (2.7) | 0.8 |
| BAL % Neutrophil, mean (SD) | 8.8 (9.6) | unknown | - |
| GAP score, mean (SD) | 3.1 (1.1) | 4.4 (1.2) | <0.01 |

IPF patients in the contemporary confirmation cohort (n=52) were older (mean age 68.7 years) and more often male (82.7%) compared to the survival confirmation cohort (Table 2). Like the survival cohort, the majority of patients in the contemporary cohort were ever-smokers (61.5%). Mean baseline FVC% was 73.2 (16.6)] with mean baseline DLCO% predicted of 42.8 (11.9). At the time of BALF collection, 42% of the contemporary cohort were taking anti-fibrotic medications and none were on immunosuppression.

### Lung tissue proteomics pathway analysis

A total of 1809 proteins were identified in at least one of the 10 tissue samples. Of these, 19 proteins were considered contaminants and excluded from analysis. Differential expression analysis of 1125 proteins detected in at least 50% of the samples yielded 70 significant proteins exhibiting at least two-fold change between the short and long survival groups (p-value < 0.05, absolute log2FC ≥1) (Figure 1A, Supplement Table 1). Of these, 69 proteins were successfully mapped to IPA canonical pathways. A total of 95 pathways were determined to be significant (p<0.05) I.E., discriminant proteins between short survival and long survival were found to be significantly enriched in these pathways. The most significant pathway based on p-value was neutrophil degranulation (p-value=8.3E-06, z-score = 1.667, number of mapped proteins = 9) (Supplement Table 2). The positive z-score = 1.667 in this analysis implies an increase in expression of the neutrophil degranulation pathway in the short survival IPF group. Selenoamino acid metabolism was ranked as the second significant pathway (p-value=2.5E-04, z-score = 0, mapped proteins = 4), and SRP-dependent co-translational protein targeting to membrane was ranked third (p-value=3.3E-04, z-score = 0, mapped proteins = 4) (Figure 1B,

**Figure 1.**
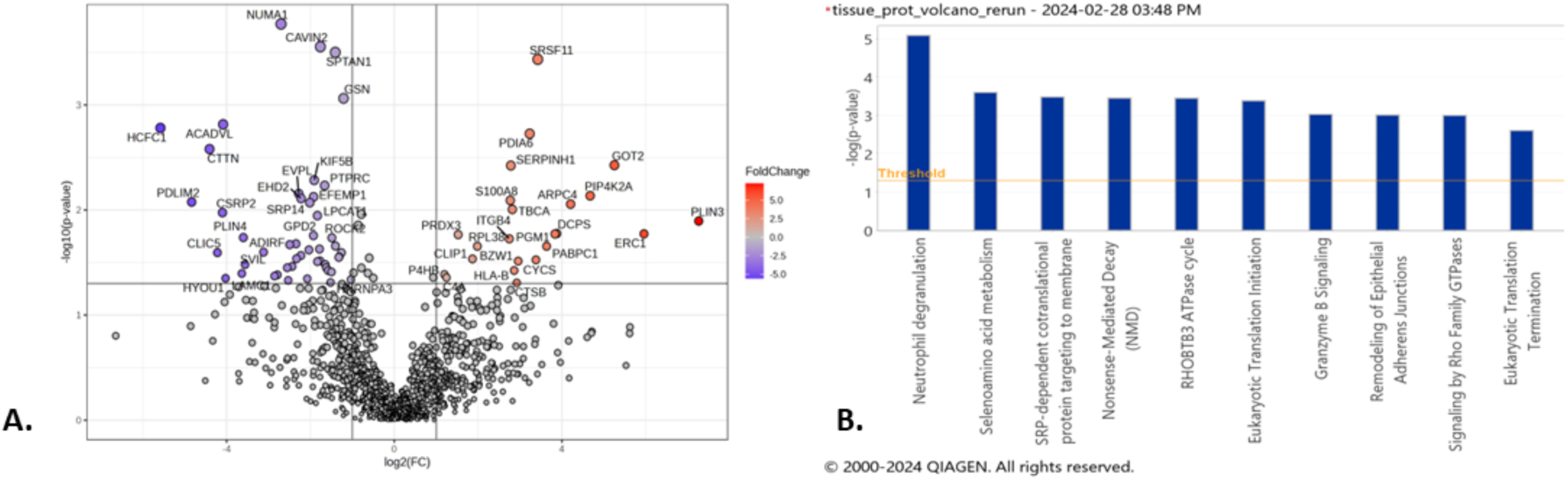
Volcano plots and canonical pathway summary graphs of differentially expressed proteins (DEP) in IPF lung tissue. A. Volcano plot of DEP in lung tissue, |log2(FC)|≥1 (2x FC in either direction), p-value < 0.05. B. IPA canonical pathway summary graph of tissue DEP showing top 10 pathways by p-value, p-value<0.05.

Supplement Table 2). The z-scores=0 of these two pathways indicated that IPA could not calculate whether they were up-regulated or down-regulated. The 9 proteins belonging to the neutrophil degranulation pathway were: CTSB, HLA-B, MVP, PDXK, PGM1, PSMA2, S100A8, SPTAN1, SRP14.

### Selection of validation target pathway

Our proteomics analyses identified neutrophil degranulation (also termed neutrophil extracellular trap formation (NETosis)) as a key pathway distinguishing different IPF survival trajectories in lung tissue, based on its prominent significance in the differential pathway analyses (smallest p-value in IPF tissue). Therefore, we elected to carry forward NETosis as the candidate pathway for confirmation in the IPF lung microenvironment using complementary absolute protein quantification methods.

### Correlation of NET markers and BALF neutrophil percentage

The percentage of BALF neutrophils significantly correlated with the levels of each BALF NET remnant (MPO-DNA: r=0.34, p<0.01; exDNA: r=0.49, p<0.01; calprotectin: r=0.51, p<0.01; NE: r=0.46, p<0.01) (Figure 2). There was also a strong positive correlation amongst the four different measures of BALF NET remnants in the survival confirmation cohort (Figure 2).

**Figure 2.**
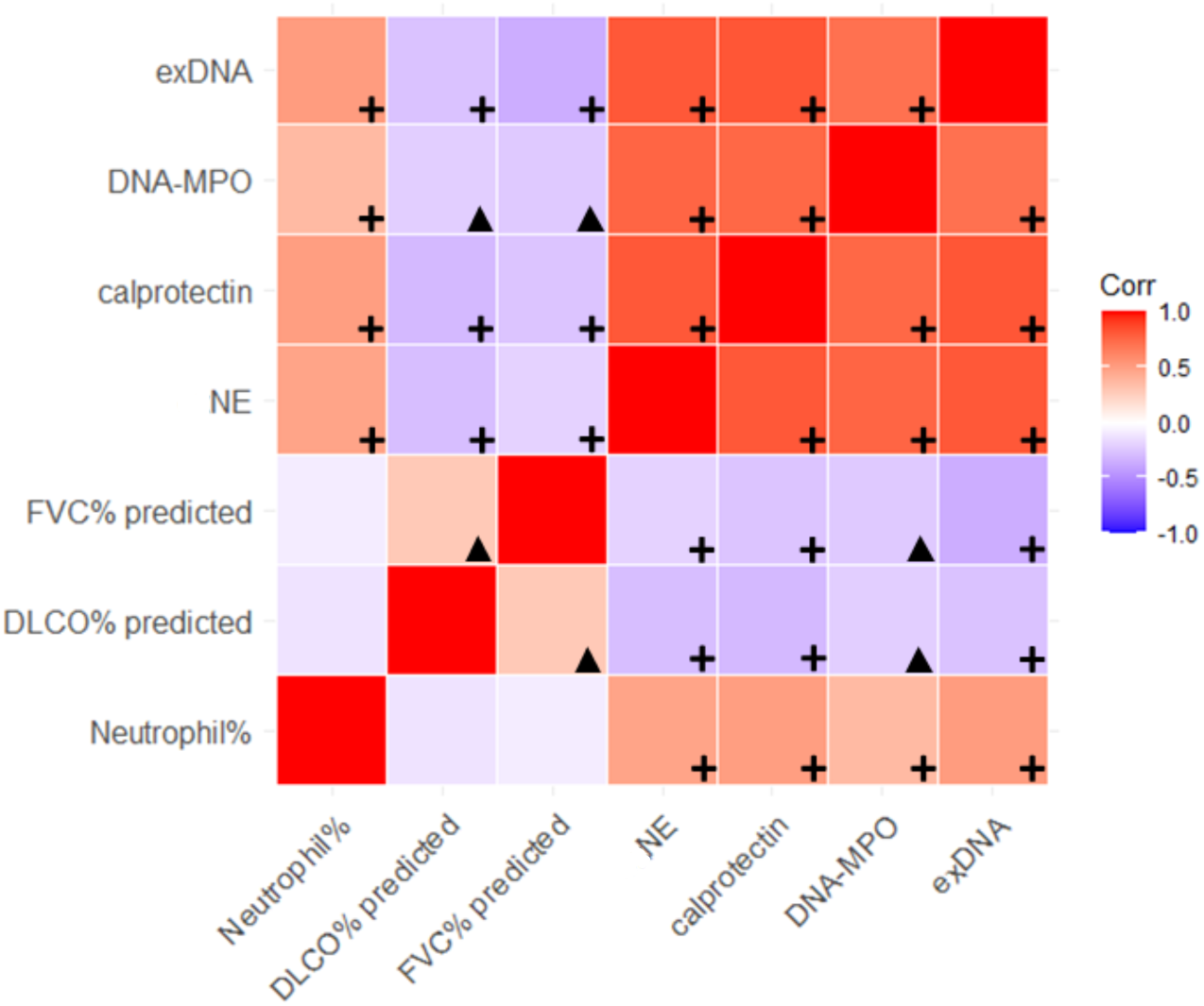
Correlogram highlighting degree of correlation across the four NET remnants, neutrophil percentage in BALF samples, and the negative correlation with FVC and DLCO (% predicted) across the discovery cohort. A red box color indicates a positive correlation with darker reds indicating greater strength of correlation and purple boxes indicate negative correlation with darker colors indicating stronger correlation, based on Pearson’s correlation coefficient. Black plus signs indicate correlations with p-value <0.01 and black triangles indicate significant correlations with p-values between 0.01 and 0.05.

### BALF NET remnant levels associated with worse lung disease severity

In the survival confirmation cohort, there was a significant negative correlation between lower FVC% predicted and higher BALF NET remnant levels [MPO-DNA: r=(-)0.19, p=0.03; exDNA: r=(-)0.34, p<0.01; calprotectin: r=(-)0.30, p<0.01; NE: r=(-)0.22, p<0.01] (Figure 3, Supplemental Figure 1). Similarly, there was a negative correlation between lower DLCO% predicted and higher BALF NET remnant levels (MPO-DNA: r=(-)0.23, p<0.01; exDNA: r=(-)0.26, p<0.01; calprotectin: r=(-)0.29, p<0.01; NE: r=(-)0.22, p<0.01) (Figure 2). Using linear regression models, each NET remnant was significantly negatively associated with FVC% after adjustment for age and sex (MPO-DNA: p=0.02; exDNA: p<0.01; calprotectin: p<0.01; NE: p<0.01) and DLCO% predicted after adjustments for age and sex (MPO-DNA: p=0.01; exDNA: p<0.01; calprotectin: p<0.01; NE: p=0.01).

**Figure 3.**
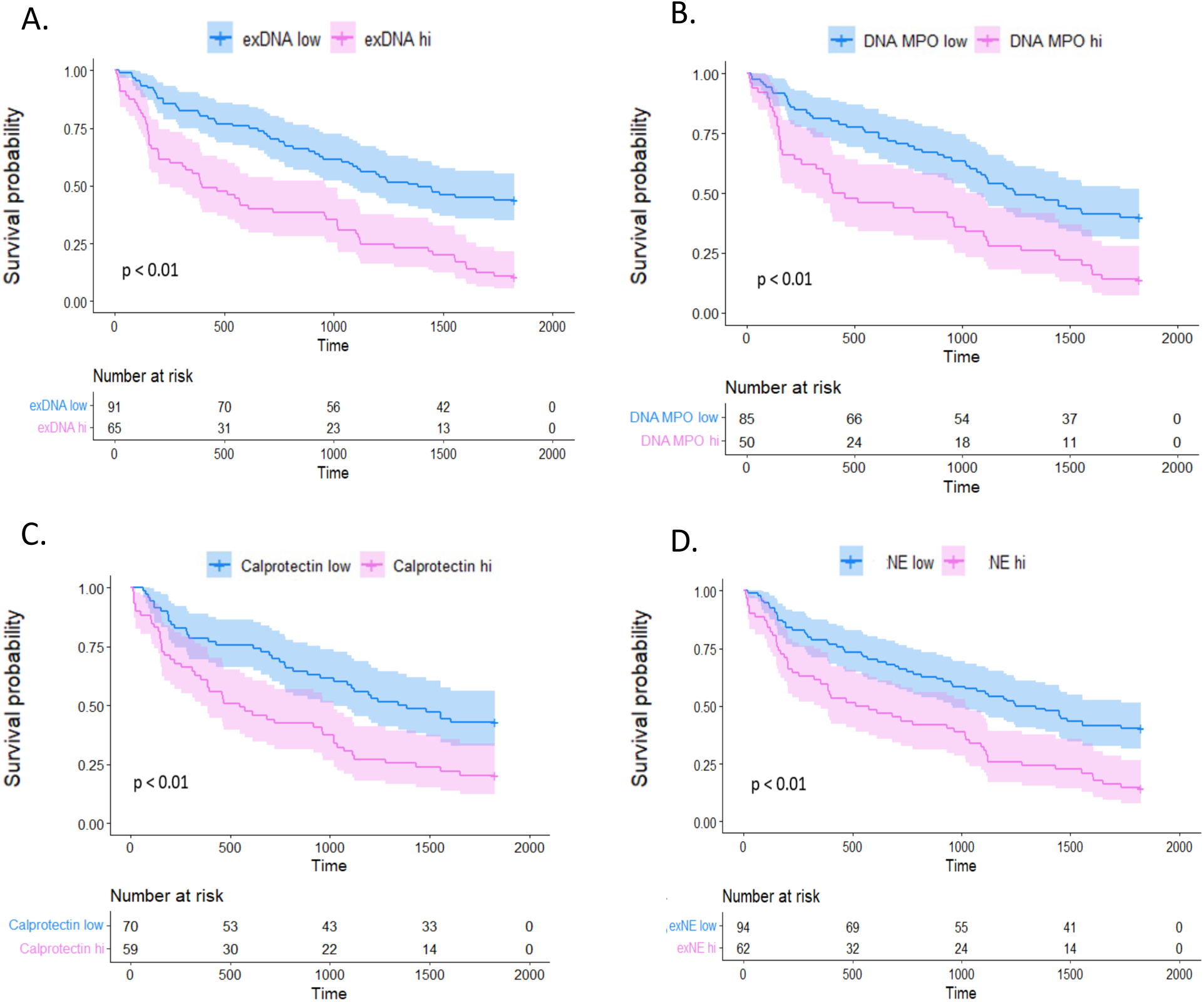
Kaplan-Meier survival curves in survival cohort. Each pane represents one of the NETosis markers tested within the validation cohort which was divided into high and low NETosis marker levels by optimal cut-off threshold. Patients with NETosis marker levels higher than the optimal cutoff threshold are represented in pink and those with NETosis marker levels below the threshold are in blue. Survival probability is represented by the y-axis and the time to death in days is plotted on the x-axis. Panel A shows results from exDNA: HR=2.57, 95% CI 1.76–3.76, p<0.01; Panel B shows results from DNA-MPO: HR=2.15, 95% CI 1.43–3.24, p<0.01; Panel C shows results from calprotectin: HR=2.00, 95% CI 1.31–3.05, p<0.01; Panel D shows results from NE: HR=2.06, 95% CI 1.41–3.01, p<0.01.

### BALF NET remnants are associated with worse survival

In the survival confirmation cohort using unadjusted models, higher levels of each of the four BALF NET remnants was associated with worse 5-year survival in the survival cohort (DNA-MPO: HR=1.19, 95% CI 1.05–1.36, p<0.01; exDNA: HR=1.32, 95% CI 1.15–1.53, p<0.01; calprotectin: HR=1.27, 95% CI 1.07–1.51, p<0.01; NE: HR=1.17, 95% CI 1.04–1.31, p=0.01). In models adjusted for GAP score and use of immunosuppression, DNA-MPO, exDNA, and calprotectin remained significantly associated with worse survival (DNA-MPO: HR=1.18, 95% CI 1.02–1.38, p<0.05; exDNA: HR=1.30, 95% CI 1.09–1.55, p<0.01; calprotectin: HR=1.23, 95% CI 1.00–1.51, p<0.05).

For each NET remnant test, IPF patients were stratified into groups based on high vs. low BALF NET levels. For all four measures of NET remnants, the high BALF NET group had significantly worse survival compared to IPF patients with low BALF NET remnant levels [DNA-MPO: HR=2.15, 95% CI 1.43–3.24, p<0.01; exDNA: HR=2.57, 95% CI 1.76–3.76, p<0.01; calprotectin: HR=2.00, 95% CI 1.31–3.05, p<0.01; NE: HR=2.06, 95% CI 1.41–3.01, p<0.01] (Figure 3). Each of these associations remained significant after adjusting for GAP score and use of immunosuppression [DNA-MPO: HR=1.98, 95% CI 1.20–3.26, p<0.01; exDNA: HR=2.19, 95% CI 1.33-3.60, p< 0.01; calprotectin: HR=1.79, 95% CI 1.05–3.03, p=0.03; NE: HR=1.82, 95% CI 1.12-2.94, p=0.01].

### Contemporary confirmation cohort

As in the survival cohort, there was a significant relationship between lower FVC% predicted and lower DLCO% predicted and higher BALF exDNA [r=(-)0.29, p=0.04 and r=(-)0.30), p=0.04, respectively] in the contemporary confirmation cohort.

## Discussion

Using untargeted label-free quantitative proteomics in IPF lung tissue, we identified NETosis as a novel pathway associated with IPF survival. We then confirmed these proteomics observations using alternative, absolute quantification methods in the IPF lung microenvironment using larger IPF cohorts. In these large IPF cohorts we observed important associations between BALF NET markers and worse baseline disease severity and worse survival. Finally, given the era of collection of our survival confirmation cohort, we confirmed the presence and importance of NETosis in the lung microenvironment of contemporary IPF patients where we again observed a relationship between BALF NET markers and worse baseline disease severity.

These data confirm the importance of NETosis in IPF via convergence of findings across two IPF lung compartments (lung tissue and BALF), using label-free proteomics followed by cross-confirmation with absolute protein quantification methodology. In this analysis we leveraged lung tissue for discovery using cutting edge system biology technology, and notably confirmed the clinical significance of NET markers in IPF BALF in two independent cohorts. Our findings in BALF are of particular interest as it is a more accessible compartment (BALF vs. lung tissue) that can allow for future NETosis focused research in IPF patients.

Previous data have highlighted the prognostic role of BALF neutrophilia and serum neutrophil-to-lymphocyte ratio for IPF patients [11, 27]. However, the mechanism by which the presence of neutrophils results in worse outcomes in IPF is unknown. Our data suggest NETosis as the possible link. NETosis occurs when neutrophils expel their DNA in complex with granule-derived proteins (Figure 4), which can contribute to the damage of surrounding tissues. These NET scaffolds play a direct role in innate immune functions to clear pathogenic debris but have also been implicated in lung injury where NET-related immunologic responses to chronic and acute inflammation are associated with worse prognosis in asthma, bronchiectasis, and acute lung injury [22, 28, 29]. NETs also induce key pathways of pulmonary fibrosis [30–33], and NET inhibition in animal models can resolve lung injury and fibrosis [30, 34]. While NETosis has been shown to have important associations with lung injury, disease severity and survival in other pulmonary conditions [22, 28, 29], this is the first in-human evidence that NETs may contribute to IPF survival and disease severity.

**Figure 4.**
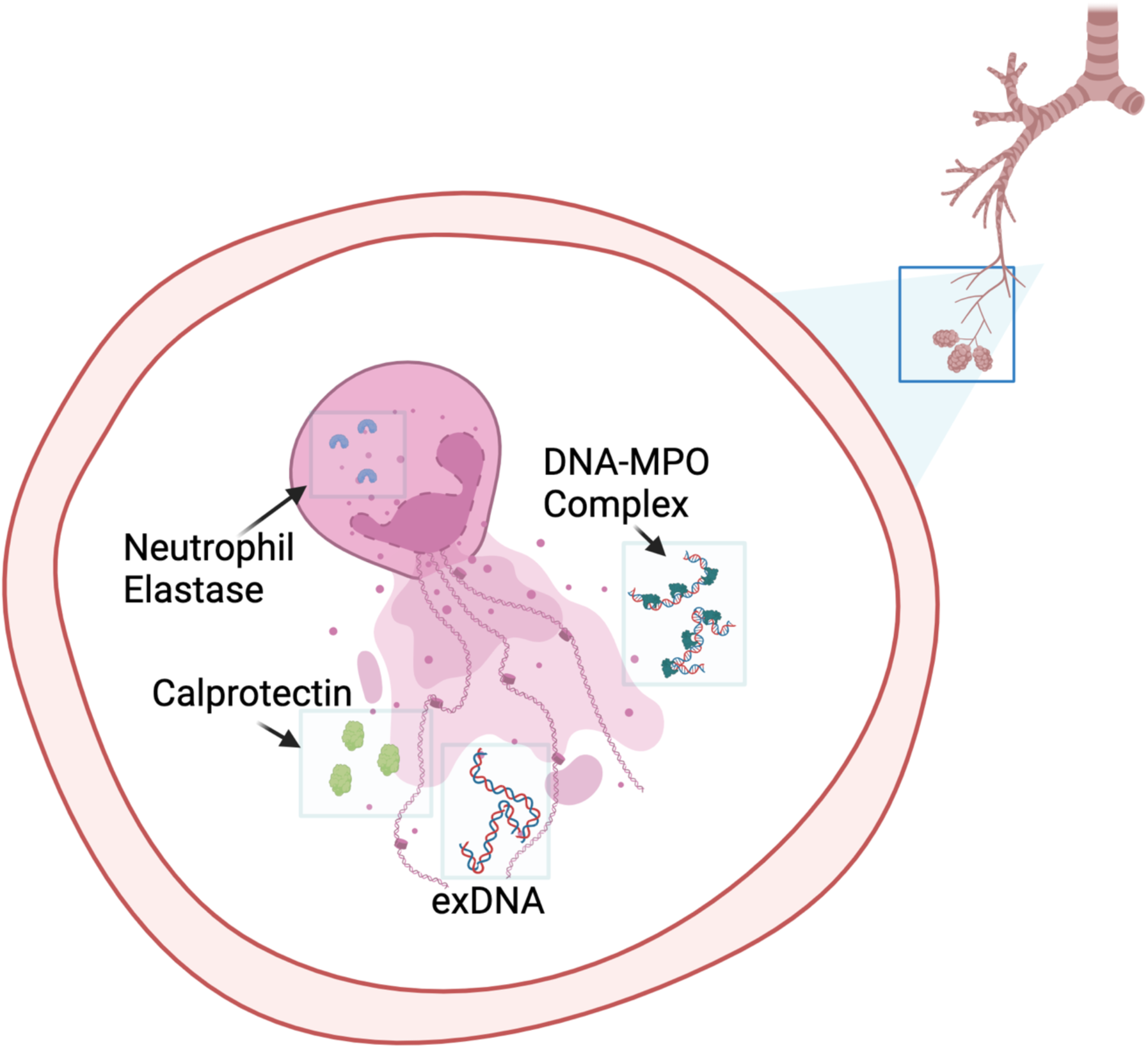
Neutrophils in the lungs after undergoing stimulation to release their intracellular protein cargo via neutrophil extracellular trap formation (NETosis). This material includes extracellular DNA (exDNA), calprotectin and DNA-myeloperoxidase (DNA-MPO) complexes. NETosis is mediated by the activity of neutrophil elastase, which is also detectable in the extracellular environment. The presence of exDNA has been associated with immunologic and inflammatory reactions that likely result in direct lung and airway injury.

There are also several interesting links between NETosis and known risk factors for IPF. Tobacco smoke and *in vitro* neutrophil stimulation by purified MUC5B induces neutrophils to undergo NETosis, and smoking and genetic variants associated with increased MUC5B production are known risk factors for IPF [35–39]. Additionally, in mouse models, inhibition of NETosis reduces bleomycin-induced lung injury [40]. Studies to date have demonstrated that some NET associated proteins can induce lung fibroblast proliferation and myofibroblast differentiation [30, 31]. Several NET proteins have also been found to be cytotoxic to alveolar epithelial cells [33]. Future studies should explore the direct effects of different NET protein cargo, including NET-derived exDNA, on lung injury in IPF.

The data from our study also offer a possible explanation for previous immunologic phenomena observed in IPF patients. There are increased circulating autoantibodies demonstrated in patients with IPF [41–44]. NETosis is associated with autoantigen exposure and the development of autoantibodies associated with diseases including rheumatoid arthritis, scleroderma, anti-neutrophil cytoplasmic antibody vasculitis, and inflammatory myositis syndromes [17, 20, 24, 45–47]. Given the role of NETs in autoantibody development, these observations also suggest that NETosis may be an important entity to better understand the complex relationship between IPF and fibrotic forms of autoimmune ILDs [48–52].

There are important considerations to interpreting these data that require future prospective study. For instance, the majority of samples included in this study were collected in a previous era of diagnosis and treatment and therefore the role of immunosuppression and anti-fibrotic medication use should be considered in these observations. For these reasons, we included a second, multi-center cohort to confirm the relationship between NETosis and disease severity in IPF lungs under contemporary practices and treatments. Additionally, because BALF is rarely obtained for clinical or research purposes in IPF in current practice, it may be challenging to incorporate these findings clinical practice.

It is important to highlight that while these data find a strong association between BALF levels of NET remnants and clinical outcomes, it remains unclear if NETosis is directly pathogenic in IPF or if it represents a marker of upstream lung injury. It is also unclear if the increased lung levels of NET remnants in some patients with IPF, are the results of neutrophils more prone to undergo NETosis or if the neutrophils in these patients encountered more factors that induced NET formation Furthermore, prospective study with uniform collection methods will be required to understand a clinically important threshold that can be validated to develop a NETosis ‘biomarker’ that could be used for assessment of prognosis and potentially inform treatment decisions in patients with IPF. Future study should also explore circulating markers of NETosis given the lack of widespread bronchoscopy in IPF clinical practice.

In conclusion, NETosis is a relatively under-explored possible mechanism of lung injury in IPF, and our data find associations between higher lung NETosis and worse clinical outcomes. Future work is needed to understand if this represents a novel target for IPF drug discovery or a pathogenic mechanism of lung injury for some IPF patients.

## Data Availability

All data produced in the present study are available upon reasonable request to the authors.

## Author contributions

SMM, JJS, and MKD contributed to the study conception and design. KKB, PJW and JJS contributed to the recruitment of the study subjects. SMM, JSL, PJW, MKD, JJS, LTN, YS, VF, CL, IA, MPW, MJR, and MKD contributed to acquisition of the data. SMM, LN, AA, EN, DK and MKD contributed to the analysis and interpretation of the data. SMM and MKD contributed to the initial drafting of the manuscript. All authors contributed to critical revision and final approval of the manuscript.

## Acknowledgements

We would like to acknowledge Hal Collard and the rest of the Weighing Risks and Benefits of Laparoscopic Anti-Reflux Surgery in Patients with IPF (WRAP-IPF) investigators for contributing samples for our validation cohort. We would also like to acknowledge Kevin K. Brown for his contribution and efforts to collect and maintain the original NJH ILD cohort.

