## Supplemental Tables for "Neutrophil extracellular traps linked to idiopathic pulmonary fibrosis severity and survival"

**Supplemental Table 1**.

| Supplement table. Differentially expressed proteins in IPF lung tissues | | | |
|---|---|---|---|
| | FC | log2(FC) | raw.pval |
| PLIN3 | 113.8 | 6.8303 | 1.34E-05 |
| HCFC1 | 0.021301 | -5.553 | 3.86E-05 |
| SRSF11 | 10.934 | 3.4507 | 0.000558 |
| GOT2 | 69.792 | 6.125 | 0.000924 |
| PIP4K2A | 28.799 | 4.848 | 0.001396 |
| PDIA6 | 11.933 | 3.5769 | 0.001752 |
| SERPINH1 | 8.6939 | 3.12 | 0.001753 |
| ERC1 | 60.984 | 5.9304 | 0.002596 |
| TBCA | 8.4263 | 3.0749 | 0.00287 |
| ARPC4 | 18.199 | 4.1858 | 0.003097 |
| CSRP2 | 0.044171 | -4.5008 | 0.003371 |
| PABPC1 | 17.825 | 4.1558 | 0.003699 |
| ACADVL | 0.076888 | -3.7011 | 0.003862 |
| PRDX3 | 3.4105 | 1.77 | 0.003918 |
| NUMA1 | 0.17715 | -2.497 | 0.004251 |
| CTTN | 0.073382 | -3.7684 | 0.004434 |
| BZW1 | 7.0332 | 2.8142 | 0.00477 |
| PDLIM2 | 0.03152 | -4.9876 | 0.005138 |
| NID2 | 0.018354 | -5.7678 | 0.005351 |
| PGM1 | 27.64 | 4.7887 | 0.006797 |
| CAVIN2 | 0.30732 | -1.7022 | 0.008871 |
| RPL38 | 5.7121 | 2.514 | 0.00984 |
| NELFB | 62.909 | 5.9752 | 0.010233 |
| PPIB | 7.6251 | 2.9308 | 0.010747 |
| S100A8 | 5.9271 | 2.5673 | 0.011707 |
| CEP170 | 11.46 | 3.5185 | 0.011912 |
| ADIRF | 0.089893 | -3.4756 | 0.012662 |
| HECTD1 | 6.5128 | 2.7033 | 0.014883 |
| DCPS | 13.537 | 3.7588 | 0.015703 |
| HINT2 | 0.16119 | -2.6331 | 0.016519 |
| SRP14 | 0.18806 | -2.4107 | 0.021065 |
| CLIC5 | 0.044844 | -4.4789 | 0.021484 |

| | | | |
|---|---|---|---|
| SPTAN1 | 0.42669 | -1.2287 | 0.022095 |
| EVPL | 0.23867 | -2.0669 | 0.022314 |
| HNMT | 0.18029 | -2.4716 | 0.022659 |
| PDXK | 8.1612 | 3.0288 | 0.023942 |
| SAA1 | 10.613 | 3.4078 | 0.025799 |
| SMTN | 0.10217 | -3.291 | 0.026498 |
| PLIN4 | 0.074885 | -3.7392 | 0.02654 |
| CGN | 0.052645 | -4.2476 | 0.02661 |
| HLA-B | 7.8322 | 2.9694 | 0.02871 |
| ITGB4 | 10.055 | 3.3298 | 0.030233 |
| HNRNPA3 | 0.39674 | -1.3337 | 0.031724 |
| LASP1 | 0.28473 | -1.8123 | 0.031765 |
| KIF5B | 0.29463 | -1.763 | 0.032184 |
| CTSB | 5.132 | 2.3595 | 0.032557 |
| HNRNPM | 4.6959 | 2.2314 | 0.033903 |
| RPS3A | 0.31807 | -1.6526 | 0.035262 |
| PURB | 0.15649 | -2.6758 | 0.035939 |
| EHD2 | 0.21471 | -2.2196 | 0.037727 |
| NME1 | 4.1509 | 2.0534 | 0.037753 |
| CLIP1 | 3.0146 | 1.592 | 0.037842 |
| MVP | 3.6004 | 1.8482 | 0.03794 |
| PSMA2 | 52.036 | 5.7014 | 0.039768 |
| CYCS | 11.512 | 3.5251 | 0.040542 |
| C4A | 2.2781 | 1.1878 | 0.040588 |
| C3(1) | 0.31969 | -1.6453 | 0.041409 |
| NPM1 | 4.2676 | 2.0934 | 0.041662 |
| GPD2 | 0.34412 | -1.539 | 0.041776 |
| TALDO1 | 2.4325 | 1.2824 | 0.042361 |
| DDRGK1 | 3.4888 | 1.8027 | 0.042875 |
| EFEMP1 | 0.23736 | -2.0749 | 0.044248 |
| ENAH | 0.22794 | -2.1333 | 0.044267 |
| CUL3 | 0.2669 | -1.9057 | 0.044506 |
| SEPTIN9 | 0.22738 | -2.1368 | 0.045621 |
| CA1 | 2.1825 | 1.126 | 0.046374 |
| CALD1 | 0.33731 | -1.5679 | 0.046733 |
| PRKAR2A | 0.69019 | -0.53493 | 0.047919 |
| USO1 | 0.36936 | -1.4369 | 0.049117 |
| RPL12 | 3.0019 | 1.5859 | 0.049173 |

Table 1. Differentially expressed proteins in IPF lung tissues with FC: fold change, raw pval = raw p value.

Supplement table 2. IPA pathway analysis summary of differentially expressed proteins in IPF lung tissue

© 2000-2024 QIAGEN. All rights reserved.

| Ingenuity Canonical Pathways | -log(p-value) | Ratio | z-score | Molecules |
|---|---|---|---|---|
| Neutrophil degranulation | 5.08 | 0.0189 | 1.667 | CTSB,HLA-B,MVP,PDXK,PGM1,PSMA2,S100A8,SPTAN1,SRP14 |
| Selenoamino acid metabolism | 3.6 | 0.0374 | 0 | HNMT,RPL12,RPL38,RPS3A |
| SRP-dependent cotranslational protein targeting to membrane | 3.48 | 0.0348 | 0 | RPL12,RPL38,RPS3A,SRP14 |
| Nonsense-Mediated Decay (NMD) | 3.45 | 0.0342 | 1 | PABPC1,RPL12,RPL38,RPS3A |
| RHOBTB3 ATPase cycle | 3.45 | 0.2 | #NUM! | CUL3,PLIN3 |
| Eukaryotic Translation Initiation | 3.38 | 0.0328 | 1 | PABPC1,RPL12,RPL38,RPS3A |
| Granzyme B Signaling | 3.03 | 0.125 | #NUM! | CYCS,NUMA1 |
